# *GCH1* genetic variation as a prognostic factor in Parkinson’s disease across populations

**DOI:** 10.64898/2026.08.14.26359677

**Authors:** Jung Hwan Shin, Maria Teresa Periñan, Joo Won Jang, Laurel Screven, Lara M. Lange, Christine Klein, Joshua M. Shulman, Ziv Gan-Or, Morvarid Ghamgosar Shahkhali, Konstantin Senkevich, Petr Dusek, Irina Miliukhina, Roy N. Alcalay, Chin-Hsien Lin, Ruey-Meei Wu, Huw R. Morris, Eng-King Tan, Bao-Rong Zhang, Guillaume Cogan, Alexis Brice, Niccolò E. Mencacci, Ignacio Juan Keller Sarmiento, Tanya Simuni, Samia Ben Sassi, M. J. Martí, Pau Pastor, Yi Wen Tay, Ai Huey Tan, Shen-Yang Lim, Maria Stamelou, Freddy Chafota, Miguel E. Rentería, Wael Mohamed, Ignacio F. Mata, Mario Cornejo Olivas, Martin Cesarini, Andrea Rivera, Micol Avenali, Enza Maria Valente, Tatiana M. Foroud, Kelly N. H. Nudelman, Michael C. Brumm, Thomas Gasser, Rimona S. Weil, Claire Shepherd, Kishore Raj Kumar, Rejko Krüger, Ken Marek, Rauan Kaiyrzhanov, Steve Gentleman, Jee-Soo Lee, Han-Joon Kim, Beomseok Jeon, the Global Parkinson’s Genetics Program (GP2)

**Author notes:** Joint first authors (Jung Hwan Shin, Maria Teresa Periñan, Joo Won Jang). **Correspondence to:** Beomseok Jeon, MD, PhD, BJ Center for comprehensive Parkinson care and rare movement disorders, Chung-Ang University Health care system, Hyundae Hospital, NamyangJu, Korea.

## Abstract

**Background:** Pathogenic variants in *GCH1* have been associated with Parkinson’s disease (PD), but the clinical phenotype and longitudinal disease course of *GCH1*-associated PD remain incompletely characterized.

**Objectives:** To characterize the genetic spectrum, clinical phenotype, and longitudinal progression of *GCH1*-associated PD across multiple populations.

**Methods:** Whole-genome sequencing (WGS) and clinical exome sequencing (CES) data from the Global Parkinson’s Genetics Program (GP2) were analyzed together with unpublished and published *GCH1-associated* PD patients. Variant pathogenicity was classified according to ACMG criteria. Demographic, clinical, and longitudinal features were compared between *GCH1* P/LP variant carriers and non-carrier PD patients; individuals with known pathogenic variants in PD-associated genes were excluded from both groups.

**Results:** In the GP2 cohort (PD, n=22,825; controls, n=4,453), 16 pathogenic or likely pathogenic (P/LP) *GCH1* variants were identified in 58 individuals, including 54 PD patients, one control, and three individuals with other neurodegenerative phenotypes (two with progressive supranuclear palsy and one with dementia with Lewy bodies). In the pooled-ancestry WGS analysis, *GCH1* P/LP variants were enriched in PD patients versus controls (0.267% vs 0.023%; OR=11.854; 95% CI=1.620-86.699; *p*=0.0006). Variant frequencies in PD patients ranged from 0.121% to 0.714% across ancestries. In the CES cohort, P/LP variants were identified in 0.201% of PD patients. After integrating GP2 with additional unpublished and published datasets, 119 *GCH1-associated* PD patients were analyzed. Compared with non-carriers, *GCH1* P/LP variant carriers had earlier disease onset (53.7 ± 14.8 vs 59.2 ± 11.7 years; *p*=8.99×10^-5^), lower levodopa equivalent daily dose requirements (467.5 ± 331.7 vs 680.3 ± 466.39mg/day; *p*=4.21×10^-8^), and more frequent family history of PD (47.6% vs 19.9%; *p*=1.09×10^-8^). Adjusted Cox models showed significant delayed progression to motor fluctuations (HR=0.32, 95% CI=0.16-0.62) and levodopa-induced dyskinesias (HR=0.51, 95% CI=0.30-0.87).

**Conclusion:** *GCH1* pathogenic variants were associated with a clinically distinct phenotype characterized by earlier disease onset and slower progression of motor complications. These findings suggest that *GCH1* genetic variants may serve as genetic biomarkers for patient stratification and prognosis in PD.

## Introduction

Heterozygous variants in GTP cyclohydrolase 1 (*GCH1*) are the major genetic cause of dopa-responsive dystonia (DRD) ^1^, a disorder characterized by childhood- or adolescent-onset dystonia, parkinsonism, and diurnal fluctuation. As DRD primarily results from dopamine deficiency rather than neurodegeneration, affected individuals typically show a marked and sustained response to low-dose levodopa ^2^. Beyond DRD, *GCH1* variants have also been associated with parkinsonism with motor fluctuations accompanied by reduced dopamine transporter (DAT) binding, consistent with presynaptic dopaminergic dysfunction and suggestive of nigrostriatal degeneration ^3–9^.

Approximately 11% of *GCH1* variant carriers develop parkinsonism in the absence of dystonia ^10^, and rare heterozygous *GCH1* variants have been reported in 0.56-1.9% of patients with PD, in whom they are significantly enriched compared with controls ^9,11–13,14,15,16^. In addition, large multi-ancestry genome-wide association studies have consistently identified *GCH1* as a PD risk locus ^17,18^. However, the clinical phenotype of *GCH1*-associated PD remains incompletely characterized. Previous studies have suggested that *GCH1*-associated PD may present with earlier disease onset and a relatively mild clinical course ^5,15,19^, but these observations have been limited by small sample sizes, cross-sectional study designs, and limited ancestral diversity. Consequently, it remains unclear whether these patients exhibit distinct clinical features compared with non-carriers.

In this study, using the multi-ancestry Global Parkinson’s Genetics Program (GP2), together with unpublished international cohorts and systematically curated published cases, we investigated the genetic and clinical spectrum of GCH1-associated PD. We aimed to characterize age at onset, longitudinal disease progression and the clinical phenotype associated with *GCH1* variants in PD.

## Methods

### Study Cohort

We analyzed whole-genome sequencing (WGS) data from GP2 Release 10 (DOI: 10.5281/zenodo.15748014), comprising 21,073 individuals, including 12,371 PD patients, 4,433 controls, and 4,269 individuals with other neurodegenerative phenotypes. Clinical exome sequencing (CES) data from 10,454 PD patients enrolled through the PD GENEration project led by the Parkinson’s Foundation and released through GP2 were also included ^24^. Because PD GENEration is a case-only cohort, controls were unavailable. Sample distributions by sequencing platform and ancestry are summarized in Supplementary Table 1.

Additional unpublished PD patients carrying *GCH1* variants from international cohorts (Canada, France, Germany, China, Australia, Japan, and Singapore) were included. Published PD patients with *GCH1* variants were identified through a PubMed search (last accessed in December 31, 2025) using the terms “GCH1” and “Parkinsonism” or “Parkinson disease” ^3–11,14,15,23,25–33^. Studies reporting individual-level genetic and clinical data were reviewed, and patients with insufficient information or suspected duplicate reports were excluded. The Institutional Review Board (IRB) of Seoul National University Hospital approved this study (IRB No. 2001-119-1097).

### Sequencing Data Processing

WGS data were processed using the Broad Institute Functional Equivalence and Joint Genotyping pipelines ^34^. High-confidence variants were retained based on call rate >95%, genotype quality > 20, read depth > 5, and heterozygous allele balance between 0.25 and 0.75. Genetic ancestry was assigned using GenoTools (https://github.com/GP2code/GenoTools) following previously described methods ^35^. Individuals were assigned to one of eleven ancestry groups: African Admixed (AAC), African (AFR), Ashkenazi Jewish (AJ), Latinos and Indigenous People of the Americas (AMR), Complex Admixture History (CAH), Central Asian (CAS), East Asian (EAS), European (EUR), Finnish (FIN), Middle Eastern (MDE), and South Asian (SAS).

### GCH1 Variant Carrier Screening and Classification

Genomic coordinates for *GCH1* were obtained from Ensembl (GRCh38: chr14: 54,842,008 - 54,902,826) (https://www.ensembl.org). Variants were extracted using PLINK v2.0 and annotated with ANNOVAR ^36,37^, using refGene, dbNSFP v4.7a, and gnomAD v4.1. Downstream analyses were restricted to coding and splicing *GCH1* variants (including missense, nonsense, frameshift, start-loss, and in-frame insertions/deletions) with minor allele frequency (MAF) <1% in gnomAD based on the overall observed frequency across all ancestral populations, and Combined Annotation Dependent Depletion (CADD) score ≥ 12.37, corresponding to the top 5-6% of predicted deleterious variants ^38^.

*GCH1* variant pathogenicity was manually classified according to the 2015 ACMG/AMP guidelines and relevant ClinGen Sequence Variant Interpretation recommendations in the context of DRD ^39^. Evidence codes and final classifications were assigned individually for each variant. Publicly available variant resources, including ClinVar and HGMD, together with ACMG/AMP-based interpretation tools (Franklin and VarSome), were reviewed as supporting evidence when applicable; however, final classifications were based on manual curation. When family data were available, segregation analyses were performed to assess co-segregation of candidate variants with disease status among affected and unaffected relatives.

### Retrospective Collection of Clinical Information

Clinical information was collected for individuals carrying pathogenic or likely pathogenic (P/LP) *GCH1* variants using a standardized case report form (CRF) (Supplementary Material). Data included demographic characteristics (age at evaluation, sex, age at last follow-up, and family history of PD), motor features (age at onset [AAO] and motor complications, including motor fluctuations, levodopa-induced dyskinesia, and freezing of gait), and non-motor features (dementia, orthostatic hypotension, urinary incontinence, and REM sleep behavior disorder). Results of DAT imaging and the presence of additional PD-associated genetic variants were also collected.

For comparisons with non-carriers, demographic and clinical data were obtained from GP2 WGS, CES, and NeuroBooster Array (NBA) datasets. To minimize confounding by other monogenic forms of PD ^40^, individuals harboring P/LP variants in established or emerging PD-associated genes, including *SNCA, LRRK2, VPS35, PRKN, PINK1, DJ-1 (PARK7), GBA1, ATP13A2, DNAJC6, FBXO7, VPS13C, SYNJ1, RAB39B, CHCHD2, DCTN1, WDR45, SLC20A2, RAB32,* and *JAM2*, were excluded from the non-carrier group. NBA data were used exclusively for non-carriers to maximize statistical power, whereas identification of *GCH1* variant carriers was restricted to sequencing-based datasets, which provide higher variant resolution and enable reliable identification of rare *GCH1* variants. Clinical measures for the non-carriers were obtained from the GP2 data release, including censoring information for time-to-event analyses of levodopa-induced dyskinesia, motor fluctuations, and dementia.

### Statistical Analysis

Continuous variables are presented as mean ± standard deviation (SD). Group comparisons were performed using Welch *t* test for continuous variables and χ^2^ test for categorical variables. Linear and logistic regression models adjusted for age, sex, and disease duration were used where appropriate.

Kaplan–Meier survival analyses were used to visualize time to clinical milestones, including motor fluctuations, levodopa-induced dyskinesia, and dementia. Time-to-event was calculated from disease onset to the occurrence of each outcome, with patients censored at their last follow-up if the event had not occurred. Survival curves were compared using the log-rank test. Median time-to-event was estimated from the Kaplan-Meier survival curves and defined as the time point at which 50% of patients had experienced the corresponding milestone. Associations between *GCH1* carrier status and the hazard of clinical milestones were evaluated using Cox proportional hazards regression models adjusted for sex and AAO. Hazard ratios (HRs) with 95% confidence intervals (CIs) and corresponding p-values were reported. Statistical analyses were performed using Python in Jupyter Notebooks.

## Results

### Genetic Characterization of GCH1 Variant Carriers

*GCH1* variants identified across the GP2 WGS and CES datasets are summarized in Supplementary Table 2. Rare heterozygous *GCH1* variants were identified in 99 individuals, representing 42 unique variants, including 86 patients with PD, 6 controls, and 7 individuals with other neurodegenerative phenotypes. Among the 86 PD patients, 54 were identified through WGS and 34 through CES, with 2 individuals present in both datasets.

Following ACMG-based interpretation, two variants (p.Pro23Leu and p.Pro69Leu) were classified as benign and excluded from downstream analyses. Of the remaining variants, 16 were classified as P/LP (Table 1) and 26 as variants of uncertain significance (VUS). P/LP *GCH1* variants were identified in 54 PD patients, 1 control, and 3 individuals with other neurodegenerative phenotypes, including progressive supranuclear palsy (PSP; n=2) and dementia with Lewy bodies (DLB; n=1).

**Table 1.**
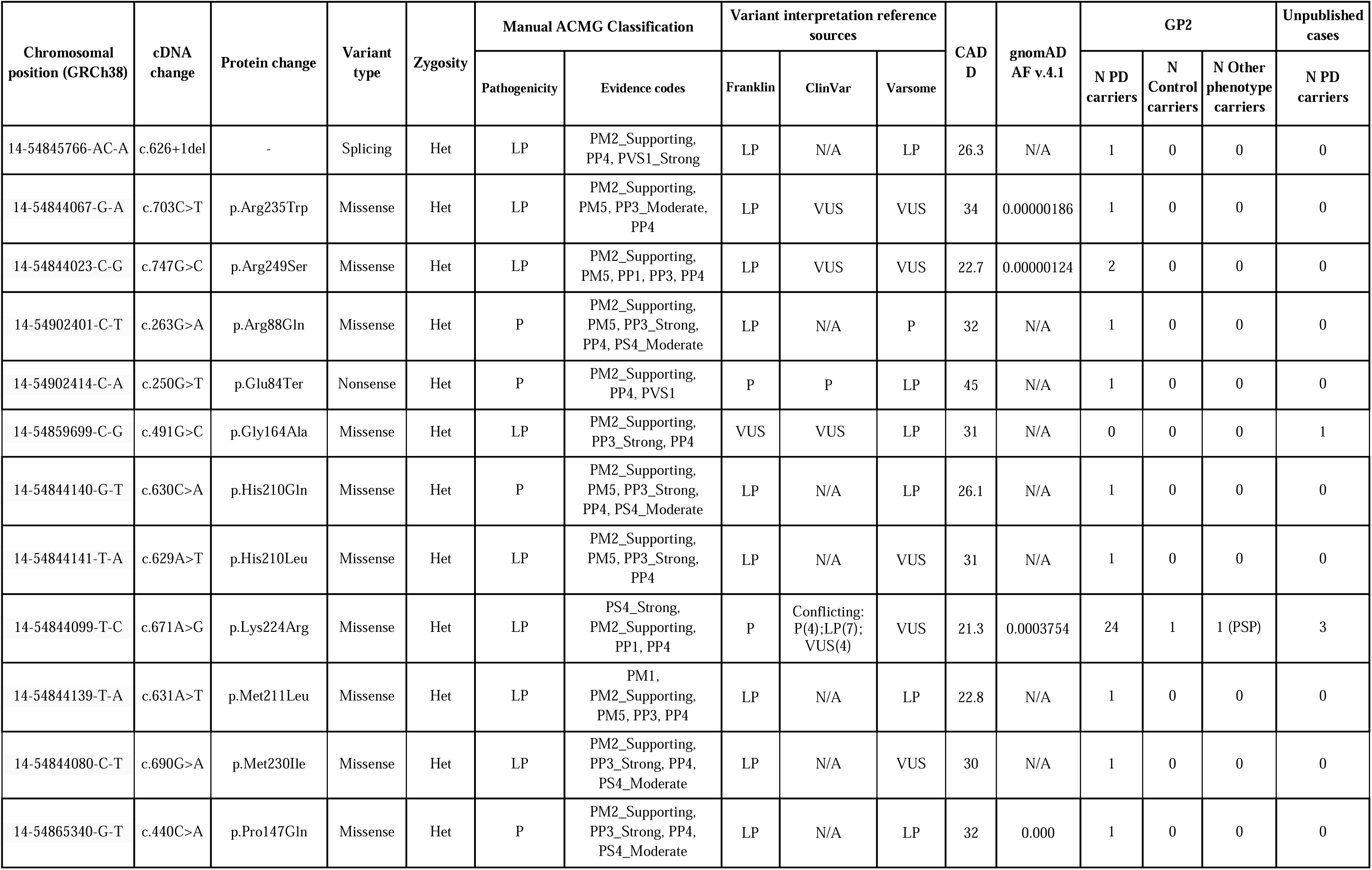
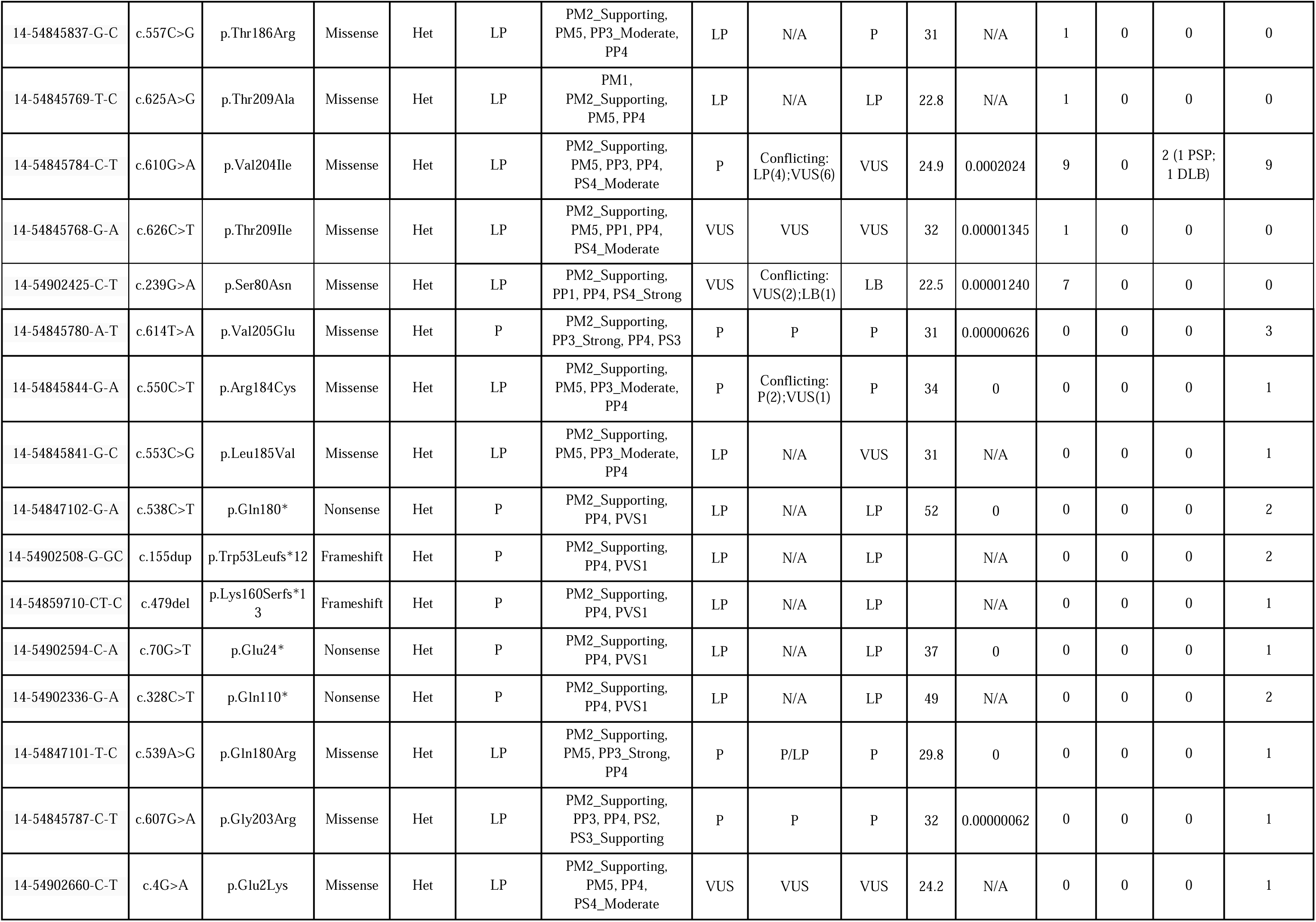
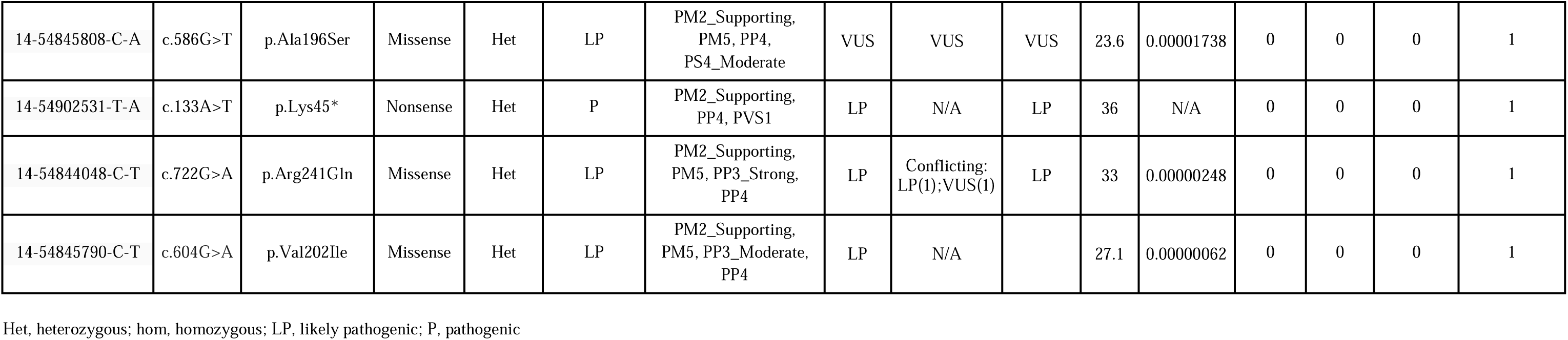
Summary of *GCH1* pathogenic or likely pathogenic variants identified in the GP2 study cohort and the unpublished cases.

Among the 54 patients with PD carrying P/LP *GCH1* variants identified across the WGS and CES datasets, 10 carried additional pathogenic variants in established PD-associated genes and were excluded from the genotype–phenotype analyses.

The recurrent p.Lys224Arg variant was identified in three affected individuals from two unrelated families (Figure 1). Segregation analysis demonstrated co-segregation with PD in one family. In the second family, the variant was present only in the affected proband and absent in two unaffected relatives carrying the co-occurring *GBA1* p.Arg296Gln variant.

**Figure 1.**
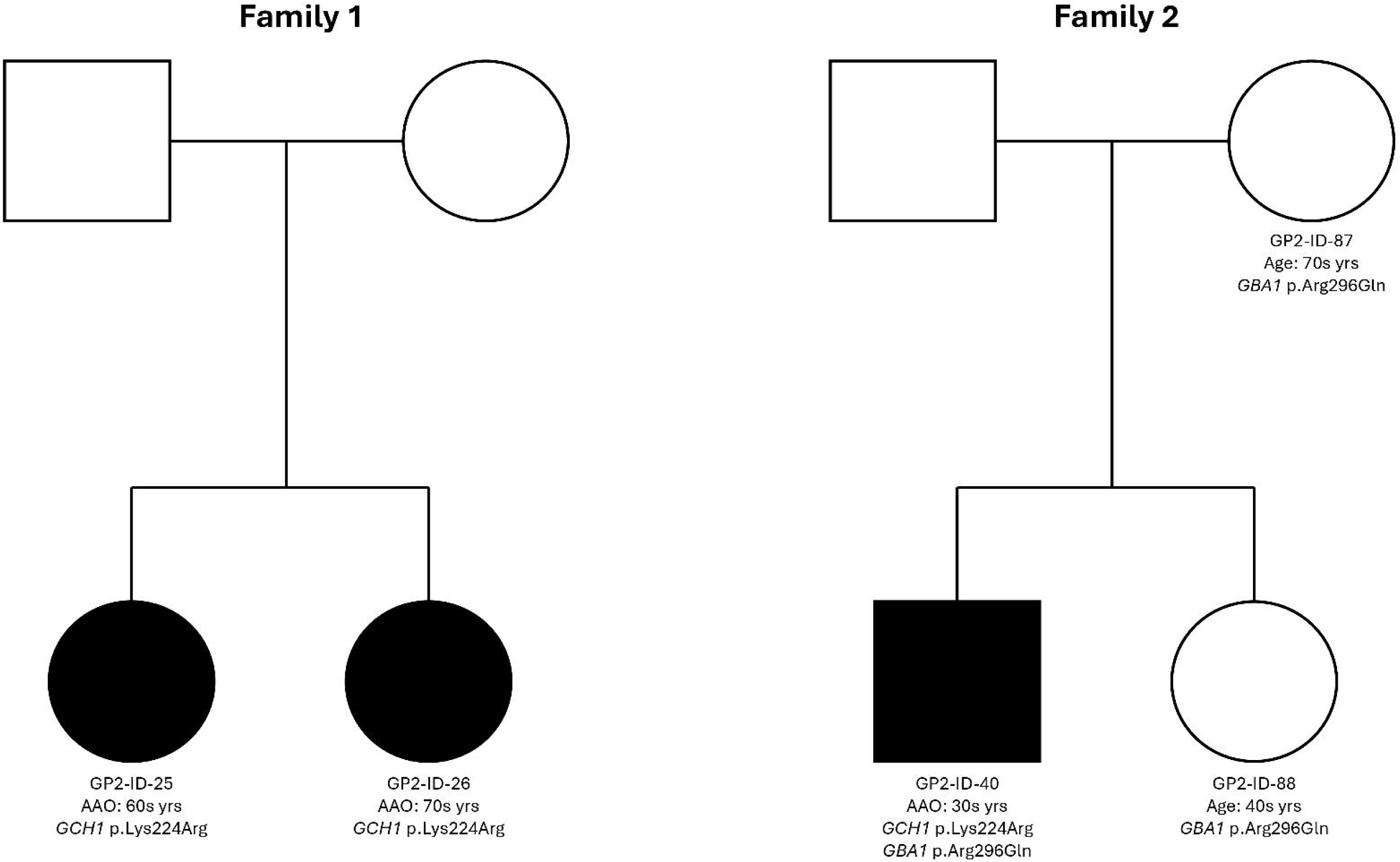
Pedigrees of two unrelated families with heterozygous *GCH1* p.Lys224Arg variants. In Family 1, two affected siblings developed Parkinson’s disease (PD) at ages in their late 50s and early 70s years, respectively, and both carried the *GCH1* p.Lys224Arg variant; clinical and genetic information on the parents were unavailable. In Family 2, the proband (GP2-ID-40) presented with early-onset PD and carried both *GCH1* p.Lys224Arg and *GBA1* p.Arg296Gln variants. The unaffected mother (GP2-ID-88) and unaffected sibling (GP2-ID-88) each carried the *GBA1* p.Arg296Gln variant alone. Filled symbols denote individuals affected by PD.

### Frequency of GCH1 Variants in PD and Controls

Case-control analyses were restricted to the GP2 WGS dataset. Rare *GCH1* variants were identified in 54 of 12,371 patients with PD and 6 of 4,433 controls. In the pooled multi-ancestry analysis, *GCH1* variants were significantly enriched in PD patients compared with controls (54/12,371 [0.437%] vs 6/4,433 [0.135%]; OR=3.235, 95% CI=1.391 - 7.524, *p*=0.0029). Restricting the analysis to P/LP variants strengthened the association (33/12,371 [0.267%] vs 1/4,433 [0.023%]; OR=11.854, 95% CI=1.620 - 86.699, *p*=0.0006). Among individuals of European ancestry, P/LP *GCH1* variants were more frequent in PD patients than controls (20/7,288 [0.274%] vs 1/1,718 [0.058%]; OR=4.725, 95% CI=0.634 - 35.231, *p*=0.158); however, this comparison did not reach statistical significance. P/LP *GCH1* variants were also identified in PD patients of East Asian (8/1,856 [0.431%]), African Admixed (1/140 [0.714%]), Central Asian (1/350 [0.286%]), Middle Eastern (2/488 [0.410%]), and African (1/826 [0.121%]) ancestries. No P/LP *GCH1* variant carriers were observed among non-European controls (n=2,715), limiting ancestry-specific case-control comparisons outside the European cohort.

In the CES dataset, which included only patients with PD, *GCH1* variants were identified in 34 of 10,454 PD patients (0.325%), including 21 (0.201%) carrying P/LP *GCH1* variants. Among these carriers, frequencies were 0.237% (11/4,641) in individuals of European ancestry, 0.376% (1/266) in American Admixed individuals, and 0.862% (1/116) in African Admixed individuals; ancestry information was unavailable for 8 carriers.

After stratifying the GP2 cohort, we included 28 unpublished and 47 previously reported patients with PD carrying P/LP GCH1 variants in the clinical phenotype analysis. The final pooled clinical cohort comprised 119 patients carrying P/LP *GCH1* variants, including 44 from GP2, 28 unpublished patients, and 47 previously published cases.

### Clinical Characteristics of PD Associated with P/LP GCH1 Variants

Clinical characteristics of *GCH1-associated* PD patients, with comparisons to PD patients without *GCH1* variants, are summarized in Table 2. *GCH1-associated* PD patients had a significantly earlier AAO than non-carrier PD patients (53.7 ± 14.8 years, *n=*112, vs 59.2 ± 11.7 years, *n*=25,062; *p*=8.99 x 10^-5^; Figure 2). A positive family history of PD was more common among *GCH1* P/LP variant carriers than non-carriers (39/82 [47.6%] vs 5,001/25,062 [19.9%]; *p*=1.09 x 10^-8^).

**Figure 2.**
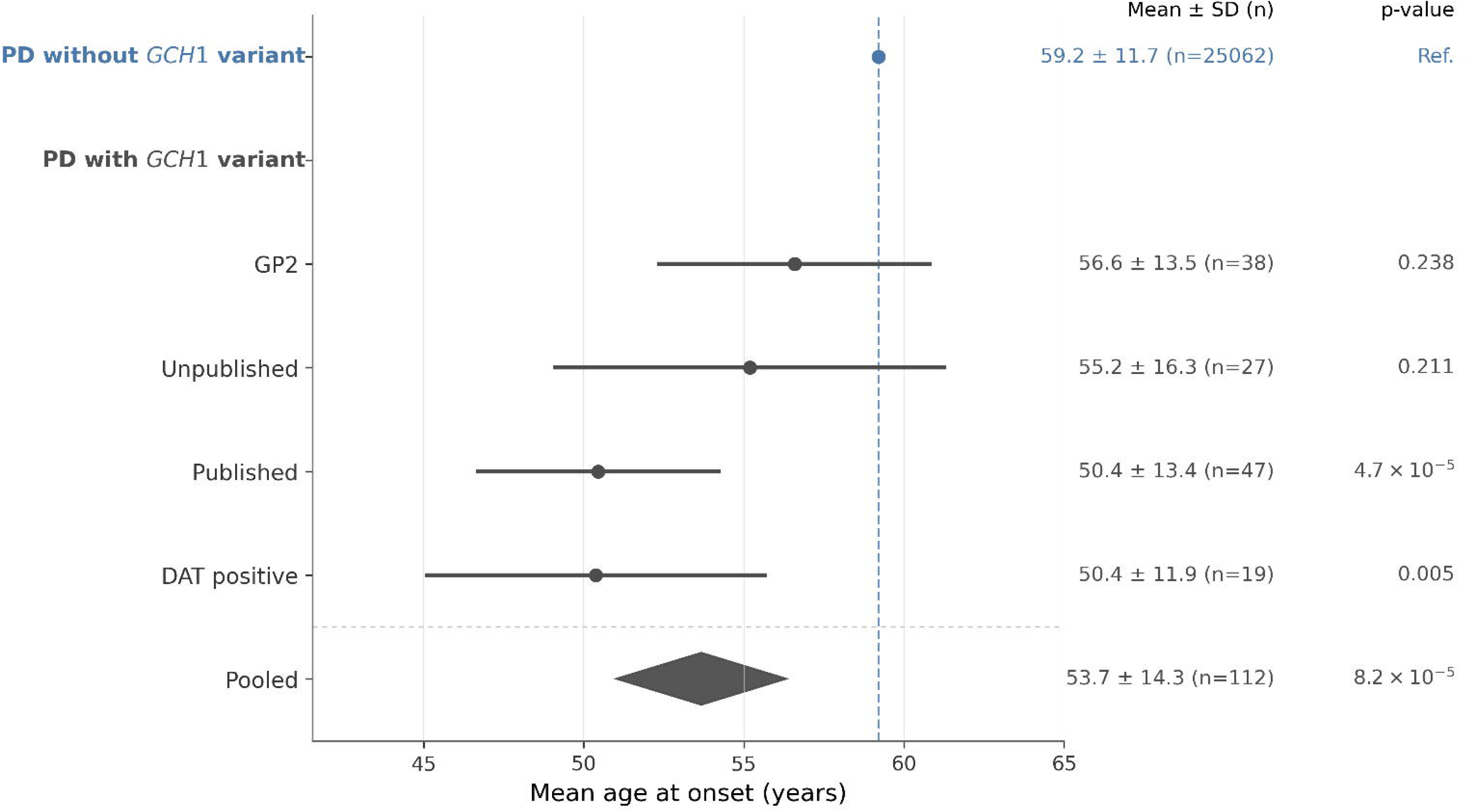
Mean age at onset (AAO) of Parkinson’s disease (PD) among individuals carrying *GCH1* variants compared with reference PD cases without *GCH1* variants. Subgroups are shown for GP2, unpublished cases, published cases, and DAT-positive cases. Circles indicate the mean AAO, and horizontal lines represent the 95% confidence intervals. The pooled estimate across all *GCH1* variant carriers is shown as a diamond. Individuals with *GCH1* variants had an earlier mean AAO than the reference PD population (53.7 ± 14.3 vs. 59.2 ± 11.7 years). Mean ± SD and sample size (n) are provided for each group, and p-values are calculated relative to the reference PD population without *GCH1* variants. DAT, dopamine transporter imaging; GP2, Global Parkinson’s Genetics Program.

**Table 2.**
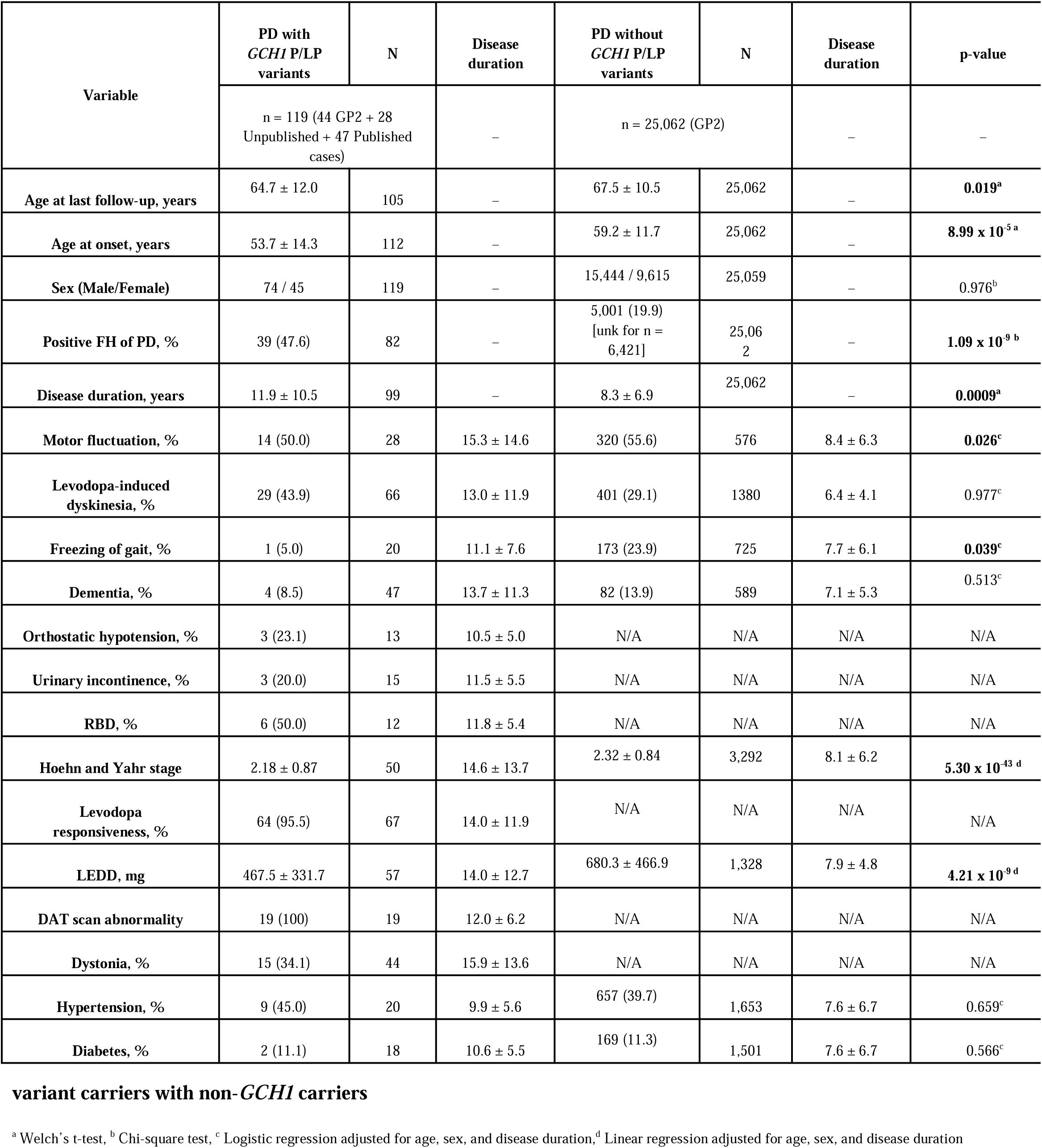
Demographic and clinical characteristics in patients with Parkinson’s disease comparing *GCH1* P/LP variant carriers with non-*GCH1* carriers.

| Variable | PD with<br>*GCH1* P/LP<br>variants | N | Disease<br>duration | PD without<br>*GCH1* P/LP<br>variants | N | Disease<br>duration | p-value |
| --- | --- | --- | --- | --- | --- | --- | --- |
|  | n = 119 (44 GP2 + 28<br>Unpublished + 47 Published<br>cases) |  | – | n = 25,062 (GP2) |  | – | – |
| Age at last follow-up, years | 64.7 ± 12.0 | 105 | – | 67.5 ± 10.5 | 25,062 | – | **0.019<sup>a</sup>** |
| Age at onset, years | 53.7 ± 14.3 | 112 | – | 59.2 ± 11.7 | 25,062 | – | **8.99 x 10<sup>-5</sup> <sup>a</sup>** |
| Sex (Male/Female) | 74 / 45 | 119 | – | 15,444 / 9,615 | 25,059 | – | 0.976<sup>b</sup> |
| Positive FH of PD, % | 39 (47.6) | 82 | – | 5,001 (19.9)<br>[unk for n =<br>6,421] | 25,062 | – | **1.09 x 10<sup>-9</sup> <sup>b</sup>** |
| Disease duration, years | 11.9 ± 10.5 | 99 | – | 8.3 ± 6.9 | 25,062 | – | **0.0009<sup>a</sup>** |
| Motor fluctuation, % | 14 (50.0) | 28 | 15.3 ± 14.6 | 320 (55.6) | 576 | 8.4 ± 6.3 | **0.026<sup>c</sup>** |
| Levodopa-induced<br>dyskinesia, % | 29 (43.9) | 66 | 13.0 ± 11.9 | 401 (29.1) | 1380 | 6.4 ± 4.1 | 0.977<sup>c</sup> |
| Freezing of gait, % | 1 (5.0) | 20 | 11.1 ± 7.6 | 173 (23.9) | 725 | 7.7 ± 6.1 | **0.039<sup>c</sup>** |
| Dementia, % | 4 (8.5) | 47 | 13.7 ± 11.3 | 82 (13.9) | 589 | 7.1 ± 5.3 | 0.513<sup>c</sup> |
| Orthostatic hypotension, % | 3 (23.1) | 13 | 10.5 ± 5.0 | N/A | N/A | N/A | N/A |
| Urinary incontinence, % | 3 (20.0) | 15 | 11.5 ± 5.5 | N/A | N/A | N/A | N/A |
| RBD, % | 6 (50.0) | 12 | 11.8 ± 5.4 | N/A | N/A | N/A | N/A |
| Hoehn and Yahr stage | 2.18 ± 0.87 | 50 | 14.6 ± 13.7 | 2.32 ± 0.84 | 3,292 | 8.1 ± 6.2 | **5.30 x 10<sup>-43</sup> <sup>d</sup>** |
| Levodopa<br>responsiveness, % | 64 (95.5) | 67 | 14.0 ± 11.9 | N/A | N/A | N/A | N/A |
| LEDD, mg | 467.5 ± 331.7 | 57 | 14.0 ± 12.7 | 680.3 ± 466.9 | 1,328 | 7.9 ± 4.8 | **4.21 x 10<sup>-9</sup> <sup>d</sup>** |
| DAT scan abnormality | 19 (100) | 19 | 12.0 ± 6.2 | N/A | N/A | N/A | N/A |
| Dystonia, % | 15 (34.1) | 44 | 15.9 ± 13.6 | N/A | N/A | N/A | N/A |
| Hypertension, % | 9 (45.0) | 20 | 9.9 ± 5.6 | 657 (39.7) | 1,653 | 7.6 ± 6.7 | 0.659<sup>c</sup> |
| Diabetes, % | 2 (11.1) | 18 | 10.6 ± 5.5 | 169 (11.3) | 1,501 | 7.6 ± 6.7 | 0.566<sup>c</sup> |

<sup>a</sup> Welch's t-test, <sup>b</sup> Chi-square test, <sup>c</sup> Logistic regression adjusted for age, sex, and disease duration, <sup>d</sup> Linear regression adjusted for age, sex, and disease duration

Despite longer disease duration, carriers required significantly lower levodopa equivalent daily doses (LEDD) after adjustment for age, sex, and disease duration. Data on levodopa responsiveness were available for 67 *GCH1* P/LP variant carriers, of whom 64 (95.5%) showed a favorable response to treatment. Motor severity, assessed using the Hoehn and Yahr scale, was significantly lower after adjustment. DAT imaging was available for 19 carriers, all of whom had abnormal DAT binding. Dystonia was reported in 15 of 44 carriers (34.1%).Motor fluctuations and freezing of gait were significantly less frequent after adjustment, whereas the prevalence of dyskinesia, dementia, hypertension, and diabetes did not differ significantly between groups.

### Longitudinal Clinical Progression of PD Associated with GCH1 Variants

Kaplan-Meier analyses demonstrated delayed progression to motor fluctuations and levodopa-induced dyskinesia among *GCH1* P/LP variant carriers (Figure 3). Median time to motor fluctuations was 12.0 years compared with 7.9 years in non-carriers, while median time to levodopa-induced dyskinesia was 17.0 versus 10.3 years, respectively (Figure 3A, B). These associations remained significant after adjustment for AAO and sex (motor fluctuations: HR, 0.32; 95% CI, 0.16-0.62; levodopa-induced dyskinesia: HR, 0.51; 95% CI 0.30-0.87). Although dementia developed later in carriers in unadjusted analyses (Figure 3C), this association was no longer significant after adjustment (HR, 0.36; 95% CI, 0.08-1.55).

**Figure 3.**
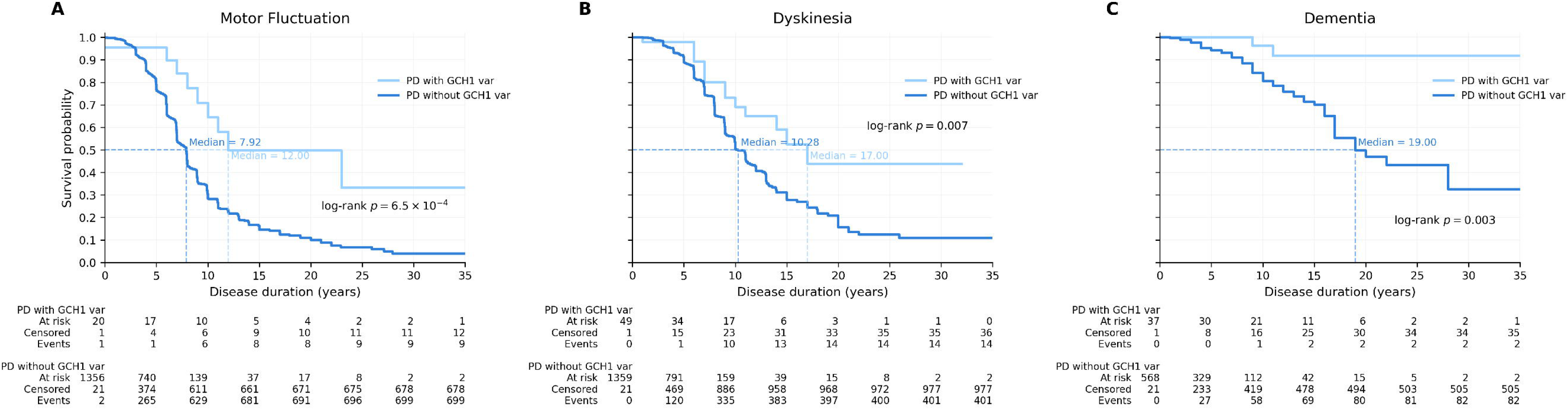
Kaplan-Meier analyses for the cumulative probability of developing key clinical milestones in Parkinson’s disease (PD) patients with and without *GCH1* variants. **(A) Time to onset of motor fluctuations. (B) Time to development of levodopa-induced dyskinesia. (C) Time to onset of dementia**. PD patients carrying *GCH1* variants exhibited later progression to motor fluctuations, dyskinesia, and dementia compared with non-carriers. Median time to each outcome is indicated by dashed lines. Group differences were assessed using log-rank tests, with corresponding p-values shown in each panel. Numbers at risk and cumulative events are provided below each plot.

## Discussion

Using the largest clinically characterized cohort of patients carrying pathogenic *GCH1* variants to date, we demonstrate that *GCH1*-associated PD is characterized by earlier disease onset and delayed progression to motor fluctuations and levodopa-induced dyskinesia. Together, these findings provide the first longitudinal evidence that *GCH1*-associated PD follows a clinical course distinct from idiopathic PD.

### Clinical Phenotype of GCH1-associated PD

Across multiple cohorts, *GCH1*-associated PD was consistently associated with an earlier age at onset, in line with previous reports^15,23^. Although statistical significance was not reached in all subgroup analyses, the direction and magnitude of the effect were consistent (Figure 2), particularly among DAT-positive patients. Phenotypic variability arising from incomplete penetrance and environmental or polygenic modifiers may partly explain the observed inter-cohort heterogeneity.

A family history of parkinsonism was substantially more common among carriers of P/LP *GCH1* variants than among non-carriers. Among carriers with available family history data, 69.2% had a family history of DRD, and 33.3% (13/39) had both DRD and parkinsonism within the same family, supporting previous observations that these phenotypes frequently coexist in *GCH1*-associated families ^20^. Additionally, nearly all carriers with available clinical data showed an excellent response to levodopa (95.5%, 64/67), and all patients with available DAT imaging showed abnormal DAT binding, consistent with previous reports and supporting the presence of presynaptic nigrostriatal degeneration rather than a purely dopamine synthesis–deficient phenotype.

An additional observation was that 14 individuals carrying concomitant pathogenic variants in established PD-associated genes (*GBA1*, *LRRK2*, *PRKN*, or *VPS13C*) were excluded from the analyses. Although the difference was not statistically significant, these individuals had a younger mean age at onset (48.6 ± 15.9 years) than the overall *GCH1* cohort (53.7 ± 14.8 years), raising the possibility that co-occurring PD-associated variants may further modify disease onset and potentially the clinical course in *GCH1*-associated PD.

### Slower Clinical Progression in GCH1-associated PD

Despite longer disease duration at the last clinical evaluation, carriers of P/LP *GCH1* variant required lower LEDD and had lower frequencies of motor fluctuations and freezing of gait than non-carriers (Table 2). These findings suggest that *GCH1-associated* PD is characterized by earlier age at onset but a comparatively milder clinical course. This contrasts with the typical expectation that patients with earlier-onset PD and longer disease duration are more likely to develop motor complications and long-term disability over time ^41^.

To account for differences in disease duration, we performed Kaplan–Meier analyses to estimate the time to clinically relevant disease milestones. Carriers of P/LP GCH1 variants had significantly longer times to the onset of motor fluctuations and levodopa-induced dyskinesia, and these associations remained significant after adjustment for age at onset (AAO) and sex in Cox proportional hazards models. Collectively, these findings indicate that *GCH1*-associated PD follows a more favourable clinical course with slower progression to motor complications. The slower disease progression was unlikely to be explained by established prognostic modifiers. The prevalence of RBD (50.0%) and orthostatic hypotension (23.1%) among P/LP *GCH1* variant carriers was comparable to that reported in idiopathic PD (∼50% and ∼30%, respectively) ^42,43,44^, and vascular risk factors, including diabetes and hypertension, did not differ significantly between groups. These findings suggest that the favourable clinical course associated with *GCH1* variants is unlikely to be explained by these factors alone.

### Mechanistic considerations

The mechanisms linking *GCH1* variation to PD remain incompletely understood. Although tetrahydrobiopterin (BH4) has been implicated in antioxidant defense, ferroptosis suppression, and mitochondrial homeostasis^45–47^, clinical observations argue against a model in which dopamine or BH4 deficiency alone is sufficient to drive progressive nigrostriatal degeneration. Experimental and clinical observations suggest that dopamine deficiency alone may be insufficient to induce progressive dopaminergic neuronal degeneration ^48^. DRD patients showed preserved nigrostriatal integrity despite decades of untreated dopamine deficiency ^48,49^. Moreover, the absence of a clear genotype-phenotype gradient linking the severity of *GCH1* dysfunction with neurodegeneration argues against a simple dose-dependent neurotoxic mechanism. Early-onset DRD, which is associated with more severe dopamine deficiency, does not typically progress to PD. Typical early-onset degenerative PD is generally associated with earlier development of dyskinesia and motor fluctuations ^50,51^. However, despite younger age at onset, GCH1-associated PD showed delayed progression to dyskinesia and motor fluctuations, suggesting a biological course distinct from typical early-onset degenerative PD.

Instead, our findings support a model in which GCH1 variants reduce dopaminergic reserve, thereby lowering the threshold for clinical manifestation of underlying nigrostriatal degeneration rather than directly causing neurodegeneration. This hypothesis is supported by reports of patients who initially presented with a DRD phenotype but subsequently developed abnormal DAT imaging and typical PD features^23^. A similar concept has been proposed for drug-induced parkinsonism, in which reduced dopaminergic reserve may unmask underlying Lewy body pathology ^52,53^. Although GCH1-associated PD clinically resembles monogenic early-onset PD caused by PINK1, PARK7, or PRKN, these genes primarily disrupt mitochondrial quality control and related neurodegenerative pathways.^54^ In contrast, our findings suggest that GCH1 primarily modifies disease expression by reducing dopaminergic reserve rather than acting as a direct driver of neurodegeneration.

### Limitations

Several limitations should be acknowledged. First, the retrospective design resulted in incomplete clinical data, and differences in phenotypic assessment across cohorts may have influenced comparisons between carriers and non-carriers. Second, the mechanistic interpretations remain speculative and require validation through longitudinal biomarker studies and experimental models. Third, DAT imaging was unavailable for many *GCH1* carriers, precluding exclusion of DRD-related parkinsonism in all cases. Fourth, GP2 cohorts may be enriched for familial or genetically predisposed PD, potentially limiting generalizability to sporadic PD populations. Consistent with this, the prevalence of family history among non-carriers was slightly higher than that reported in typical idiopathic PD cohorts ^55^. Finally, the lower frequency of P/LP *GCH1* variants observed in GP2 (0.267% in WGS and 0.201% in CES) compared with previous reports (0.46%–1.9%)^9,11,14,15^, likely reflects our conservative ACMG-based variant classification strategy and the absence of systematic copy number variant analysis.

### Conclusions

In conclusion, PD associated with pathogenic *GCH1* variants represents a genetically defined subgroup characterized c by earlier disease onset and slower progression to motor complications. These findings extend the role of *GCH1* beyond genetic susceptibility and suggest that *GCH1* status may have value for prognostic stratification and future precision medicine approaches in PD. Further longitudinal biomarker and experimental studies are needed to clarify the biological mechanisms underlying these observations.

## Supporting information

Supplementary Figure 1, Supplementary Table 1-2 and case report form

## Data Availability

Data used in the preparation of this article were obtained from the Global Parkinson Genetics Program (https://gp2.org). Specifically, we used Tier 2 data from GP2 release 10 (10.5281/zenodo.15748014). GP2 data can be accessed through AMP PD (https://amp-pd.org).
All code generated for this article, and the identifiers for all software programs and packages used, are available on GitHub (https://github.com/GP2code/) and were given a persistent identifier via Zenodo (DOI 10.5281/zenodo.21879880).

https://doi.org/10.5281/zenodo.21879880

## Acknowledgment

This project was supported by the Global Parkinson’s Genetics Program (GP2; https://gp2.org). GP2 is funded by the Aligning Science Across Parkinson’s (ASAP) (https://ror.org/03zj4c476) initiative and implemented by The Michael J. Fox Foundation for Parkinson’s Research (MJFF) (https://ror.org/03arq3225). For a complete list of GP2 members, see doi.org/10.5281/zenodo.7904831. We also acknowledge the GEoPD Consortium (https://www.geopd.net/) for its support of this work.

Some of the biospecimens used in the analyses and included in this manuscript were obtained from the Northwestern University Movement Disorders Center (MDC) Biorepository. As such, the investigators within MDC Biorepository contributed to the design and implementation of the MDC Biorepository and/or provided data and collected biospecimens but may not have participated in the analysis or writing of this manuscript. MDC Biorepository investigators include Rizwan Akhtar, MD, PhD; Tanya Simuni, MD; Dimitri Krainc, MD, PhD; Puneet Opal, MD, PhD; Joanna Blackburn, MD; and Monika Szela, MHA. A gift from the Malkin family generously supported the work of the MDC Biorepository. Part of the data used in the preparation of this article were obtained from the Canadian Open Parkinson Network (C-OPN) database (www.COPN-RPCO.ca). This Project has been made possible by Brain Canada through the Canada Brain Research Fund, with financial support of Health Canada and Parkinson Canada

## Data and Code Availability

Data used in the preparation of this article were obtained from the Global Parkinson’s Genetics Program (GP2; https://gp2.org). Specifically, we used Tier 2 data from GP2 release 10 (10.5281/zenodo.15748014). GP2 data can be accessed through AMP PD (https://amp-pd.org).

All code generated for this article, and the identifiers for all software programs and packages used, are available on GitHub (https://github.com/GP2code/) and were given a persistent identifier via Zenodo (DOI: 10.5281/zenodo.21879880).

## Author contributions

Concept and design: Jung Hwan Shin, Beomseok Jeon.

Acquisition of the data: All authors

Analysis, or interpretation of data: Jung Hwan Shin, Maria Teresa Periñan, Joo Won Jang

Drafting of the manuscript: Jung Hwan Shin, Maria Teresa Periñan, Joo Won Jang

Critical revision of the manuscript for important intellectual content: All authors.

Statistical analysis: Jung Hwan Shin, Maria Teresa Periñan, Joo Won Jang.

Supervision: Beomseok Jeon

## References

1. Segawa M, Nomura Y, Yukishita S, Nishiyama N, Yokochi M. Is phenotypic variation of hereditary progressive dystonia with marked diurnal fluctuation/dopa-responsive dystonia (HPD/DRD) caused by the difference of the locus of mutation on the GTP cyclohydrolase 1 (GCH-1) gene? Adv Neurol. 2004;94:217–223.

2. Wijemanne S, Jankovic J. Dopa-responsive dystonia--clinical and genetic heterogeneity. Nat Rev Neurol. 2015;11(7):414–424.

3. Kikuchi A, Takeda A, Fujihara K, et al. Arg(184)His mutant GTP cyclohydrolase I, causing recessive hyperphenylalaninemia, is responsible for dopa-responsive dystonia with parkinsonism: a case report. Mov Disord. 2004;19(5):590–593.

4. Tassin J, Dürr A, Bonnet AM, et al. Levodopa-responsive dystonia. GTP cyclohydrolase I or parkin mutations? Brain. 2000;123 ( Pt 6):1112–1121.

5. Lewthwaite AJ, Lambert TD, Rolfe EB, et al. Novel GCH1 variant in Dopa-responsive dystonia and Parkinson’s disease. Parkinsonism Relat Disord. 2015;21(4):394–397.

6. Eggers C, Volk AE, Kahraman D, et al. Are Dopa-responsive dystonia and Parkinson’s disease related disorders? A case report. Parkinsonism Relat Disord. 2012;18(5):666–668.

7. Hjermind LE, Johannsen LG, Blau N, et al. Dopa-responsive dystonia and early-onset Parkinson’s disease in a patient with GTP cyclohydrolase I deficiency? Mov Disord. 2006;21(5):679–682.

8. Ceravolo R, Nicoletti V, Garavaglia B, Reale C, Kiferle L, Bonuccelli U. Expanding the clinical phenotype of DYT5 mutations: is multiple system atrophy a possible one? Neurology. 2013;81(3):301–302.

9. Mencacci NE, Isaias IU, Reich MM, et al. Parkinson’s disease in GTP cyclohydrolase 1 mutation carriers. Brain. 2014;137(Pt 9):2480–2492.

10. Weissbach A, Pauly MG, Herzog R, et al. Relationship of Genotype, Phenotype, and Treatment in Dopa-Responsive Dystonia: MDSGene Review. Mov Disord. 2022;37(2):237–252.

11. Xu Q, Li K, Sun Q, et al. Rare GCH1 heterozygous variants contributing to Parkinson’s disease. Brain. 2017;140(7):e41.

12. Lange LM, Fang ZH, Screven L, et al. Rare but Relevant? Assessing Variants in Dystonia-Linked Genes in Parkinson’s Disease. Mov Disord. 2026;41(1):247–259.

13. Westenberger A, Skrahina V, Usnich T, et al. Relevance of genetic testing in the gene-targeted trial era: the Rostock Parkinson’s disease study. Brain. 2024;147(8):2652–2667.

14. Guella I, Sherman HE, Appel-Cresswell S, Rajput A, Rajput AH, Farrer MJ. Parkinsonism in GTP cyclohydrolase 1 mutation carriers. Brain. 2015;138(Pt 5):e349.

15. Yoshino H, Nishioka K, Li Y, et al. GCH1 mutations in dopa-responsive dystonia and Parkinson’s disease. J Neurol. 2018;265(8):1860–1870.

16. Boura I, Sait S, Marinakis NM, et al. The genetic architecture of Parkinson’s disease on the Island of Crete. NPJ Parkinsons Dis. 2025;11(1):354.

17. The Global Parkinson’s Genetics Program (GP2), Leonard HL. Novel Parkinson’s Disease Genetic Risk Factors Within and Across European Populations. medRxiv. Published online March 17, 2025:2025.03.14.24319455. doi:10.1101/2025.03.14.24319455

18. Kim JJ, Vitale D, Otani DV, et al. Multi-ancestry genome-wide association meta-analysis of Parkinson’s disease. Nature Genetics. 2023;56(1):27–36.

19. Pan HX, Zhao YW, Mei JP, et al. GCH1 variants contribute to the risk and earlier age-at-onset of Parkinson’s disease: a two-cohort case-control study. Transl Neurodegener. 2020;9(1):31.

20. Nygaard TG, Takahashi H, Heiman GA, Snow BJ, Fahn S, Calne DB. Long-term treatment response and fluorodopa positron emission tomographic scanning of parkinsonism in a family with dopa-responsive dystonia. Ann Neurol. 1992;32(5):603–608.

21. Jeon BS, Jeong JM, Park SS, et al. Dopamine transporter density measured by [123I]beta-CIT single-photon emission computed tomography is normal in dopa-responsive dystonia. Ann Neurol. 1998;43(6):792–800.

22. Uladzislau Rudakou, Bouchra Ouled Amar Bencheikh, Jennifer A Ruskey, Lynne Krohn, Sandra B Laurent, Dan Spiegelman, Christopher Liong, Stanley Fahn, Cheryl Waters, Oury Monchi, Edward A Fon, Yves Dauvilliers, Roy N Alcalay, Nicolas Dupré, Ziv Gan-Or. Common and rare GCH1 variants are associated with Parkinson’s disease. Neurobiol Aging. 2019;73:231.e1–e231.e6.

23. Shin JH, Lee WW, Lee JY, Kim HJ, Jeon B. GCH-1 genetic variant may cause Parkinsonism by unmasking the subclinical nigral pathology. J Neurol. 2020;267(7):1952–1959.

24. Cook L, Verbrugge J, Schwantes-An TH, et al. Parkinson’s disease variant detection and disclosure: PD GENEration, a North American study. Brain. 2024;147(8):2668–2679.

25. Hoenicka J, Vidal L, Godoy M, Ochoa JJ, García de Yébenes J. New nonsense mutation in the GTP-cyclohydrolase I gene in L-DOPA responsive dystonia-parkinsonism. Mov Disord. 2001;16(2):364–366.

26. Uncini A, De Angelis MV, Di Fulvio P, et al. Wide expressivity variation and high but no gender-related penetrance in two dopa-responsive dystonia families with a novel GCH-I mutation. Mov Disord. 2004;19(10):1139–1145.

27. Momma K, Funayama M, Li Y, et al. A new mutation in the GCH1 gene presents as early-onset Parkinsonism. Parkinsonism Relat Disord. 2009;15(2):160–161.

28. Irie S, Kanazawa N, Ryoh M, Mochizuki H, Nomura Y, Segawa M. A case of parkinsonism and dopa-induced severe dyskinesia associated with novel mutation in the GTP cyclohydrolase I gene. Parkinsonism Relat Disord. 2011;17(10):769–770.

29. Wu-Chou YH, Yeh TH, Wang CY, et al. High frequency of multiexonic deletion of the GCH1 gene in a Taiwanese cohort of dopa-response dystonia. Am J Med Genet B Neuropsychiatr Genet. 2010;153B(4):903–908.

30. Dobričić V, Tomić A, Branković V, et al. GCH1 mutations are common in Serbian patients with dystonia-parkinsonism: Challenging previously reported prevalence rates of DOPA-responsive dystonia. Parkinsonism Relat Disord. 2017;45:81–84.

31. Yan YP, Zhang B, Shen T, et al. Study of GCH1 and TH genes in Chinese patients with Parkinson’s disease. Neurobiol Aging. 2018;68:159.e3–e159.e6.

32. Sun ZF, Zhang YH, Guo JF, et al. Genetic diagnosis of two dopa-responsive dystonia families by exome sequencing. PLoS One. 2014;9(9):e106388.

33. Bernal-Pacheco O, Oyama G, Briton A, et al. A Novel DYT-5 Mutation with Phenotypic Variability within a Colombian Family. Tremor Other Hyperkinet Mov (N Y). 2013;3. doi:10.7916/D86W98SW

34. Regier AA, Farjoun Y, Larson DE, et al. Functional equivalence of genome sequencing analysis pipelines enables harmonized variant calling across human genetics projects. Nat Commun. 2018;9(1):4038.

35. Vitale D, Koretsky MJ, Kuznetsov N, et al. GenoTools: an open-source Python package for efficient genotype data quality control and analysis. G3 (Bethesda). 2025;15(1). doi:10.1093/g3journal/jkae268

36. Chang CC, Chow CC, Tellier LC, Vattikuti S, Purcell SM, Lee JJ. Second-generation PLINK: rising to the challenge of larger and richer datasets. Gigascience. 2015;4(1):s13742-015 - 0047-0048

37. Wang K, Li M, Hakonarson H. ANNOVAR: functional annotation of genetic variants from high-throughput sequencing data. Nucleic Acids Res. 2010;38(16):e164.

38. Kircher M, Witten DM, Jain P, O’Roak BJ, Cooper GM, Shendure J. A general framework for estimating the relative pathogenicity of human genetic variants. Nat Genet. 2014;46(3):310–315.

39. Richards S, Aziz N, Bale S, et al. Standards and guidelines for the interpretation of sequence variants: a joint consensus recommendation of the American College of Medical Genetics and Genomics and the Association for Molecular Pathology. Genet Med. 2015;17(5):405–424.

40. Morris H, Lim SY. Monogenic Parkinson Disease Overview. In: Adam MP, Bick S, Mirzaa GM, Pagon RA, Wallace SE, Amemiya A, eds. GeneReviews. University of Washington, Seattle; 2004.

41. Shin J, Park SH, Shin C, et al. Submandibular gland is a suitable site for alpha synuclein pathology in Parkinson disease. Parkinsonism Relat Disord. 2019;58:35–39.

42. Postuma RB, Gagnon JF, Bertrand JA, Génier Marchand D, Montplaisir JY. Parkinson risk in idiopathic REM sleep behavior disorder: preparing for neuroprotective trials. Neurology. 2015;84(11):1104–1113.

43. Velseboer DC, de Haan RJ, Wieling W, Goldstein DS, de Bie RMA. Prevalence of orthostatic hypotension in Parkinson’s disease: a systematic review and meta-analysis. Parkinsonism Relat Disord. 2011;17(10):724–729.

44. Senard JM, Raï S, Lapeyre-Mestre M, et al. Prevalence of orthostatic hypotension in Parkinson’s disease. J Neurol Neurosurg Psychiatry. 1997;63(5):584–589.

45. Kraft VAN, Bezjian CT, Pfeiffer S, et al. GTP cyclohydrolase 1/tetrahydrobiopterin counteract ferroptosis through lipid remodeling. ACS Cent Sci. 2020;6(1):41–53.

46. Soula M, Weber RA, Zilka O, et al. Metabolic determinants of cancer cell sensitivity to canonical ferroptosis inducers. Nat Chem Biol. 2020;16(12):1351–1360.

47. Tang D, Chen X, Kang R, Kroemer G. Ferroptosis: molecular mechanisms and health implications. Cell Res. 2021;31(2):107–125.

48. Rose SJ, Harrast P, Donsante C, et al. Parkinsonism without dopamine neuron degeneration in aged l-dopa-responsive dystonia knockin mice. Mov Disord. 2017;32(12):1694–1700.

49. Segawa M, Nomura Y, Hayashi M. Dopa-responsive dystonia is caused by particular impairment of nigrostriatal dopamine neurons different from those involved in Parkinson disease: evidence observed in studies on Segawa disease. Neuropediatrics. 2013;44(2):61–66.

50. Kostic V, Przedborski S, Flaster E, Sternic N. Early development of levodopa-induced dyskinesias and response fluctuations in young-onset Parkinson’s disease. Neurology. 1991;41(2 ( Pt 1)):202–205.

51. Schrag A, Schott JM. Epidemiological, clinical, and genetic characteristics of early-onset parkinsonism. Lancet Neurol. 2006;5(4):355–363.

52. Erro R, Bhatia KP, Tinazzi M. Parkinsonism following neuroleptic exposure: A double-hit hypothesis? Mov Disord. 2015;30(6):780–785.

53. Rajput AH, Rozdilsky B, Hornykiewicz O, Shannak K, Lee T, Seeman P. Reversible drug-induced parkinsonism. Clinicopathologic study of two cases. Arch Neurol. 1982;39(10):644–646.

54. Menon PJ, Sambin S, Criniere-Boizet B, et al. Genotype-phenotype correlation in PRKN-associated Parkinson’s disease. NPJ Parkinsons Dis. 2024;10(1):72.

55. Tran J, Anastacio H, Bardy C. Genetic predispositions of Parkinson’s disease revealed in patient-derived brain cells. NPJ Parkinsons Dis. 2020;6:8.

