## Supplementary Figure 1, Supplementary Table 1-2 and case report form for "*GCH1* genetic variation as a prognostic factor in Parkinson’s disease across populations": Supplementary Material_.docx

**Supplementary Figures**

**Supplementary Figure 1. Pedigree of Family 3 carrying the heterozygous *GCH1* p.Tyr175Phe variant.** The affected proband (GP2-ID-66) developed Parkinson’s disease (PD) in 70s and carried the *GCH1* p.Tyr175Phe variant. An unaffected offspring (GP2-ID-90) also carried the variant, while the unaffected spouse (GP2-ID-89) did not carry the variant. Filled symbols denote individuals affected by PD. AAO, age at onset.

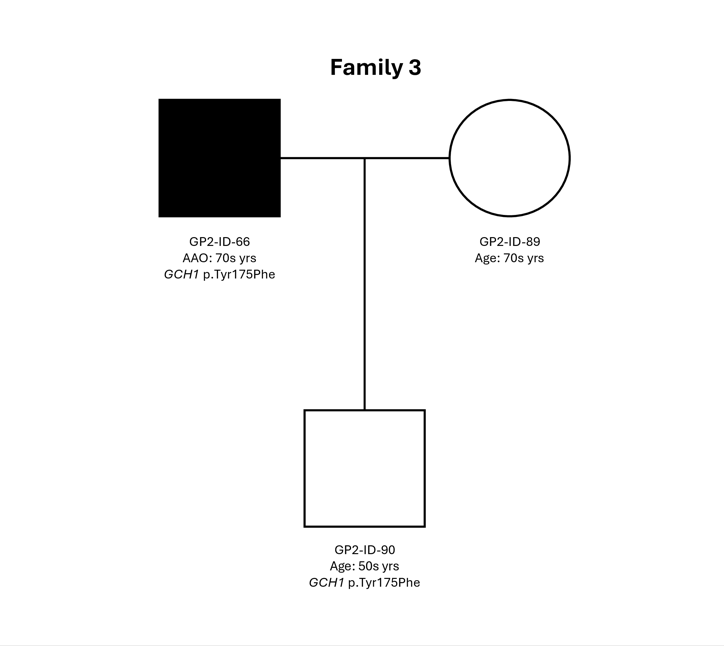

**Supplementary Tables**

**Supplementary Table 1. Demographics of the studied cohorts from GP2 Release 10 data.**

| **Genetic ancestry** | **Data type** | **Number of subjects, n** | | |
| --- | --- | --- | --- | --- |
|  |  | ***N* PD** | ***N* Other phenotypes** | ***N* Control** |
| AAC | WGS | 140 | 13 | 114 |
|  | CES | 116 | - | - |
| AFR | WGS | 826 | 16 | 854 |
|  | CES | 65 | - | - |
| AJ | WGS | 732 | 126 | 627 |
|  | CES | 611 | - | - |
| AMR | WGS | 272 | 20 | 41 |
|  | CES | 266 | - | - |
| CAH | WGS | 109 | 30 | 39 |
|  | CES | 331 | - | - |
| CAS | WGS | 350 | 157 | 333 |
|  | CES | 29 | - | - |
| EAS | WGS | 1856 | 321 | 365 |
|  | CES | 77 | - | - |
| EUR | WGS | 7288 | 3455 | 1718 |
|  | CES | 4641 | - | - |
| FIN | WGS | 22 | 4 | 4 |
|  | CES | 16 | - | - |
| MDE | WGS | 488 | 23 | 313 |
|  | CES | 40 | - | - |
| SAS | WGS | 288 | 104 | 25 |
|  | CES | 72 | - | - |
| **Total** | **WGS** | **12371** | **4269** | **4433** |
|  | **CES** | **10454**  **[ ancestry unk for n = 4190]** | **-** | **-** |

PD, Parkinson’s disease; WGS, whole-genome sequencing; CES, clinical exome sequencing

**Supplementary Table 2.** **Summary of *GCH1* variants identified in the GP2 study cohort and the unpublished cases**

| **Chromosomal position (GRCh38)** | **cDNA change** | **Protein change** | **Variant type** | **Zygosity** | **Manual ACMG classification** | | **Variant interpretation reference sources** | | | **CADD** | **gnomAD AF v.4.1** | **GP2** | | | **Unpublished cases** |
| --- | --- | --- | --- | --- | --- | --- | --- | --- | --- | --- | --- | --- | --- | --- | --- |
|  |  |  |  |  | **Pathogenicity** | **Evidence codes** | **Franklin** | **ClinVar** | **Varsome** |  |  | **N PD carriers** | **N Control carriers** | **N Other phenotype carriers** |  |
| 14-54859678-T-C | c.509+3A>G | p.? | Splicing | Het | VUS | PM2_Supporting, PP4 | VUS | Conflicting:  VUS(5);LB(2) | LB | 10.77 | 0.00006150 | 0 | 0 | 0 | 7 |
| 14-54845766-AC-A | c.626+1del | p.? | Splicing | Het | LP | PM2_Supporting, PP4, PVS1_Strong | LP | N/A | LP | 26.3 | N/A | 1 | 0 | 0 | 0 |
| 14-54902363-C-T | c.301G>A | p.Ala101Thr | Missense | Het | VUS | PM2_Supporting, PP3_Moderate, PP4 | VUS | VUS | VUS | 26.6 | 6.200e-7 | 1 | 0 | 0 | 0 |
| 14-54902642-C-A | c.22G>T | p.Ala8Ser | Missense | Het | VUS | PM2_Supporting, PP4 | VUS | VUS | LB | 12.57 | 0.00001146 | 1 | 0 | 0 | 0 |
| 14-54845801-C-T | c.593G>A | p.Arg198Gln | Missense | Het | VUS | PM2_Supporting, PM5, PP4 | VUS | VUS | VUS | 13.67 | 0.00002670 | 0 | 1 | 2 (1 PSP; 1 Prodromal) | 0 |
| 14-54902462-G-A | c.202C>T | p.Leu68Phe | Missense | Het | VUS | PM2_Supporting, PP4 | VUS | VUS | VUS | 26 | 0.000002481 | 0 | 0 | 0 | 1 |
| 14-54859699-C-G | c.491G>C | p.Gly164Ala | Missense | Het | LP | PM2_Supporting, PP3_Strong, PP4 | VUS | VUS | LP | 31 | N/A | 0 | 0 | 0 | 1 |
| 14-54845825-G-A | c.569C>T | p.Ala190Val | Missense | Het | VUS | PM2_Supporting, PP3, PP4 | LP | N/A | VUS | 31 | N/A | 0 | 0 | 0 | 1 |
| 14-54845801-C-A | c.593G>T | p.Arg198Leu | Missense | Het | VUS | PM2_Supporting, PM5, PP4 | VUS | N/A | VUS | 21.4 | N/A | 0 | 0 | 0 | 1 |
| 14-54845802-G-A | c.592C>T | p.Arg198Trp | Missense | Het | VUS | PM2_Supporting, PP3, PP4 | LP | VUS | VUS | 25.8 | 0.000005592 | 0 | 1 | 0 | 1 |
| 14-54843801-A-C | c.*216T>G | p.? | 3’UTR | Het | LB | BP4, BP7, PM2_Supporting, PP4 | N/A | N/A | LB | 8.77 | 0.000001239 | 0 | 0 | 0 | 1 |
| 14-54844067-G-A | c.703C>T | p.Arg235Trp | Missense | Het | LP | PM2_Supporting, PM5, PP3_Moderate, PP4 | LP | VUS | VUS | 34 | 0.000001859 | 1 | 0 | 0 | 0 |
| 14-54844023-C-G | c.747G>C | p.Arg249Ser | Missense | Het | LP | PM2_Supporting, PM5, PP1, PP3, PP4 | LP | VUS | VUS | 22.7 | 0.000001239 | 2 | 0 | 0 | 0 |
| 14-54844022-T-A | c.748A>T | p.Ser250Cys | Missense | Het | VUS | PM2_Supporting, PP3, PP4 | LP | VUS | VUS | 30 | 6.195e-7 | 1 | 0 | 0 | 0 |
| 14-54902489-G-C | c.175C>G | p.Arg59Gly | Missense | Het | VUS | PM2_Supporting, PP4 | VUS | VUS | VUS | 24.1 | 0.000002485 | 1 | 0 | 0 | 0 |
| 14-54902488-C-T | c.176G>A | p.Arg59His | Missense | Het | VUS | PM2_Supporting, PP4 | VUS | N/A | VUS | 24.1 | 0.000004971 | 1 | 0 | 1 (AD) | 0 |
| 14-54902401-C-T | c.263G>A | p.Arg88Gln | Missense | Het | P | PM2_Supporting, PM5, PP3_Strong, PP4, PS4_Moderate | LP | N/A | P | 32 | N/A | 1 | 0 | 0 | 0 |
| 14-54902472-G-C | c.192C>G | p.Asn64Lys | Missense | Het | VUS | PM2_Supporting, PP4 | VUS | N/A | VUS | 15.63 | 6.203e-7 | 1 | 0 | 0 | 0 |
| 14-54902454-G-T | c.210C>A | p.Asn70Lys | Missense | Het | VUS | PM2_Supporting, PP4 | VUS | N/A | VUS | 17.27 | 0.000001860 | 1 | 0 | 0 | 0 |
| 14-54844060-T-C | c.710A>G | p.Asp237Gly | Missense | Het | VUS | PM2_Supporting, PP3_Moderate, PP4 | LP | N/A | VUS | 33 | N/A | 1 | 0 | 0 | 0 |
| 14-54844052-T-A | c.718A>T | p.Thr240Ser | Missense | Het | VUS | PM2_Supporting, PP4 | VUS | N/A | VUS | 27.2 | 6.196e-7 | 0 | 0 | 0 | 1 |
| 14-54902476-T-C | c.188A>G | p.Asp63Gly | Missense | Het | VUS | PM2_Supporting, PP4 | VUS | VUS | VUS | 25.3 | N/A | 1 | 0 | 0 | 0 |
| 14-54902336-G-C | c.328C>G | p.Gln110Glu | Missense | Het | VUS | PM2_Supporting, PP4, PS4_Moderate | VUS | VUS | LP | 22.5 | 0.0001129 | 8 | 1 | 0 | 0 |
| 14-54902399-G-C | c.265C>G | p.Gln89Glu | Missense | Het | VUS | PM2_Supporting, PP4 | VUS | N/A | VUS | 22.1 | N/A | 1 | 0 | 0 | 0 |
| 14-54902414-C-A | c.250G>T | p.Glu84Ter | Nonsense | Het | P | PM2_Supporting, PP4, PVS1 | P | P | LP | 45 | N/A | 1 | 0 | 0 | 0 |
| 14-54902573-C-G | c.91G>C | p.Gly31Arg | Missense | Het | VUS | PM2_Supporting, PP4 | VUS | VUS | VUS | 19.06 | 0.000 | 1 | 0 | 0 | 0 |
| 14-54844140-G-T | c.630C>A | p.His210Gln | Missense | Het | P | PM2_Supporting, PM5, PP3_Strong, PP4, PS4_Moderate | LP | N/A | LP | 26.1 | N/A | 1 | 0 | 0 | 0 |
| 14-54844141-T-A | c.629A>T | p.His210Leu | Missense | Het | LP | PM2_Supporting, PM5, PP3_Strong, PP4 | LP | N/A | VUS | 31 | N/A | 1 | 0 | 0 | 0 |
| 14-54847120-T-C | c.520A>G | p.Ile174Val | Missense | Het | VUS | PM2_Supporting, PP4 | VUS | N/A | VUS | 22.1 | 0.000 | 1 | 0 | 0 | 0 |
| 14-54865328-T-C | c.452A>G | p.Lys151Arg | Missense | Het | VUS | PM2_Supporting, PP4 | VUS | VUS | VUS | 20.7 | 0.000 | 0 | 1 | 0 | 0 |
| 14-54844099-T-C | c.671A>G | p.Lys224Arg | Missense | Het | LP | PS4_Strong, PM2_Supporting, PP1, PP4 | P | Conflicting  :P(4);LP(7);  VUS(4) | VUS | 21.3 | 0.0003754 | 24 | 1 | 1 (PSP) | 3 |
| 14-54865395-T-C | c.385A>G | p.Met129Val | Missense | Het | VUS | PM2_Supporting, PP3_Moderate, PP4 | VUS | N/A | VUS | 23.1 | N/A | 1 | 0 | 0 | 0 |
| 14-54844139-T-A | c.631A>T | p.Met211Leu | Missense | Het | LP | PM1, PM2_Supporting, PM5, PP3, PP4 | LP | N/A | LP | 22.8 | N/A | 1 | 0 | 0 | 0 |
| 14-54844108-A-G | c.662T>C | p.Met221Thr | Missense | Het | VUS | PM2_Supporting, PP4, PS4_Moderate | LP | Conflicting  :P(1);VUS(6) | VUS | 24.1 | 0.00004152 | 2 | 0 | 0 | 1 |
| 14-54844080-C-T | c.690G>A | p.Met230Ile | Missense | Het | LP | PM2_Supporting, PP3_Strong, PP4, PS4_Moderate | LP | N/A | VUS | 30 | N/A | 1 | 0 | 0 | 0 |
| 14-54865340-G-T | c.440C>A | p.Pro147Gln | Missense | Het | P | PM2_Supporting, PP3_Strong, PP4, PS4_Moderate | LP | N/A | LP | 32 | 0.000 | 1 | 0 | 0 | 0 |
| 14-54902417-CCAGCGAGCTCAGGATGGA-C | c.229_246del | p.Ser77_Leu82del | In-frame deletion | Het | VUS | PM2_Supporting, PM4, PP4 | LP | VUS | LP | 22.4 | N/A | 1 | 0 | 0 | 0 |
| 14-54902434-G-C | c.230C>G | p.Ser77Cys | Missense | Het | VUS | PM2_Supporting, PM5, PP4 | VUS | VUS | VUS | 24.8 | 0.000001240 | 1 | 0 | 0 | 0 |
| 14-54902425-C-T | c.239G>A | p.Ser80Asn | Missense | Het | LP | PM2_Supporting, PP1, PP4, PS4_Strong | VUS | Conflicting:  VUS(2);LB(1) | LB | 22.5 | 0.00001240 | 7 | 0 | 0 | 0 |
| 14-54845837-G-C | c.557C>G | p.Thr186Arg | Missense | Het | LP | PM2_Supporting, PM5, PP3_Moderate, PP4 | LP | N/A | P | 31 | N/A | 1 | 0 | 0 | 0 |
| 14-54845769-T-C | c.625A>G | p.Thr209Ala | Missense | Het | LP | PM1, PM2_Supporting, PM5, PP4 | LP | N/A | LP | 22.8 | N/A | 1 | 0 | 0 | 0 |
| 14-54845768-G-A | c.626C>T | p.Thr209Ile | Missense | Het | LP | PM2_Supporting, PM5, PP1, PP4, PS4_Moderate | VUS | VUS | VUS | 32 | 0.00001345 | 1 | 0 | 0 | 0 |
| 14-54902506-C-A | c.158G>T | p.Trp53Leu | Missense | Het | VUS | PM2_Supporting, PP4 | VUS | VUS | VUS | 23 | 0.000001872 | 1 | 0 | 0 | 0 |
| 14-54847116-T-A | c.524A>T | p.Tyr175Phe | Missense | Het | VUS | PM2_Supporting, PM5, PP4 | VUS | VUS | VUS | 21.7 | 0.00002422 | 2 | 1 | 0 | 0 |
| 14-54845822-A-C | c.572T>G | p.Val191Gly | Missense | Het | VUS | PM2_Supporting, PM5, PP4 | LP | N/A | VUS | 23 | 0.00002111 | 1 | 0 | 0 | 0 |
| 14-54845823-C-T | c.571G>A | p.Val191Ile | Missense | Het | VUS | PM2_Supporting, PP4, PS4_Moderate | VUS | VUS | VUS | 15.22 | 0.00008323 | 2 | 0 | 1 (DLB) | 0 |
| 14-54845786-C-CCGA | c.605_607dup | p.Val202dup | In-frame duplication | Het | VUS | PM2_Supporting, PP4 | LP | N/A | LP | 18.55 | N/A | 1 | 0 | 0 | 0 |
| 14-54845784-C-T | c.610G>A | p.Val204Ile | Missense | Het | LP | PM2_Supporting, PM5, PP3, PP4, PS4_Moderate | P | Conflicting:  LP(4);VUS(6) | VUS | 24.9 | 0.0002024 | 9 | 0 | 2 (1 PSP; 1 DLB) | 9 |
| 14-54844130-C-T | c.640G>A | p.Val214Ile | Missense | Het | VUS | PM2_Supporting, PP4 | VUS | N/A | VUS | 23.5 | 6.207e-7 | 1 | 0 | 0 | 0 |
| 14-54845780-A-T | c.614T>A | p.Val205Glu | Missense | Het | P | PM2_Supporting, PP3_Strong, PP4, PS3 | P | P | P | 31 | 0.00000626 | 0 | 0 | 0 | 3 |
| 14-54845844-G-A | c.550C>T | p.Arg184Cys | Missense | Het | LP | PM2_Supporting, PM5, PP3_Moderate, PP4 | P | Conflicting: P(2);VUS(1) | P | 34 | 0 | 0 | 0 | 0 | 1 |
| 14-54845841-G-C | c.553C>G | p.Leu185Val | Missense | Het | LP | PM2_Supporting, PM5, PP3_Moderate, PP4 | LP | N/A | VUS | 31 | N/A | 0 | 0 | 0 | 1 |
| 14-54847102-G-A | c.538C>T | p.Gln180* | Nonsense | Het | P | PM2_Supporting, PP4, PVS1 | LP | N/A | LP | 52 | 0 | 0 | 0 | 0 | 2 |
| 14-54902508-G-GC | c.155dup | p.Trp53Leufs*12 | Frameshift | Het | P | PM2_Supporting, PP4, PVS1 | LP | N/A | LP | N/A | N/A | 0 | 0 | 0 | 2 |
| 14-54859710-CT-C | c.479del | p.Lys160Serfs*13 | Frameshift | Het | P | PM2_Supporting, PP4, PVS1 | LP | N/A | LP | N/A | N/A | 0 | 0 | 0 | 1 |
| 14-54902594-C-A | c.70G>T | p.Glu24* | Nonsense | Het | P | PM2_Supporting, PP4, PVS1 | LP | N/A | LP | 37 | 0 | 0 | 0 | 0 | 1 |
| 14-54902336-G-A | c.328C>T | p.Gln110* | Nonsense | Het | P | PM2_Supporting, PP4, PVS1 | LP | N/A | LP | 49 | N/A | 0 | 0 | 0 | 2 |
| 14-54847101-T-C | c.539A>G | p.Gln180Arg | Missense | Het | LP | PM2_Supporting, PM5, PP3_Strong, PP4 | P | P/LP | P | 29.8 | 0 | 0 | 0 | 0 | 1 |
| 14-54845787-C-T | c.607G>A | p.Gly203Arg | Missense | Het | LP | PM2_Supporting, PP3, PP4, PS2, PS3_Supporting | P | P | P | 32 | 0.00000062 | 0 | 0 | 0 | 1 |
| 14-54902660-C-T | c.4G>A | p.Glu2Lys | Missense | Het | LP | PM2_Supporting, PM5, PP4, PS4_Moderate | VUS | VUS | VUS | 24.2 | N/A | 0 | 0 | 0 | 1 |
| 14-54845808-C-A | c.586G>T | p.Ala196Ser | Missense | Het | LP | PM2_Supporting, PM5, PP4, PS4_Moderate | VUS | VUS | VUS | 23.6 | 0.00001738 | 0 | 0 | 0 | 1 |
| 14-54902531-T-A | c.133A>T | p.Lys45* | Nonsense | Het | P | PM2_Supporting, PP4, PVS1 | LP | N/A | LP | 36 | N/A | 0 | 0 | 0 | 1 |
| 14-54844048-C-T | c.722G>A | p.Arg241Gln | Missense | Het | LP | PM2_Supporting, PM5, PP3_Strong, PP4 | LP | Conflicting: LP(1);VUS(1) | LP | 33 | 0.00000248 | 0 | 0 | 0 | 1 |
| 14-54845790-C-T | c.604G>A | p.Val202Ile | Missense | Het | LP | PM2_Supporting, PM5, PP3_Moderate, PP4 | LP | N/A |  | 27.1 | 0.00000062 | 0 | 0 | 0 | 1 |

Het, heterozygous; hom, homozygous; LP, likely pathogenic; P, pathogenic
